# Using convolutional neural network to predict remission of diabetes after gastric bypass surgery: a machine learning study from the Scandinavian Obesity Surgery Register

**DOI:** 10.1101/2020.11.03.20224956

**Authors:** Yang Cao, Ingmar Näslund, Erik Näslund, Johan Ottosson, Scott Montgomery, Erik Stenberg

## Abstract

**Background:** Prediction of diabetes remission is an important topic in the evaluation of patients with type-2 diabetes (T2D) before bariatric surgery. While several high-quality predictive indices are available, artificial intelligence (AI) algorithms offer the potential for higher predictive capability. The objective was to construct and validate an AI prediction model for diabetes remission after Roux-en-Y gastric bypass surgery.

**Methods:** Patients who underwent surgery from 2007 until 2017 were included in the study, with collection of individual data from the Scandinavian Obesity Surgery Registry (SOReg), the Swedish National Patients Register, the Swedish Prescribed Drugs Register, and Statistics Sweden. A 7-layer convolution neural network (CNN) model was developed using 80% of patients randomly selected from SOReg and 20% of patients for external testing. The predictive capability of the CNN model and currently used scores (DiaRem, Ad-DiaRem, DiaBetter and IMS) were compared.

**Results:** In total, 8057 patients with T2D were included in the study. At 2 years after surgery 77.1% achieved pharmacological remission, while 62.2% achieved complete remission. The area under the receiver operating curve (AUC) for the CNN-model for pharmacological remission was 0.85 [95% confidence interval (CI): 0.83-0.86] during validation, and 0.83 for the final test, which was 9-12% better than the traditional predictive indices. AUC for complete remission was 0.83 (95% CI: 0.81-0.85) during validation, and 0.82 for the final test, which was 9-11% better than the traditional predictive indices.

**Conclusion:** The CNN method had better predictive capability compared to traditional indices for diabetes remission. However, further validation is needed in other countries to evaluate its external generalizability.

## 1. Introduction

Bariatric surgery is an efficient and safe treatment for patients with morbid obesity and type-2 diabetes (T2D) [1, 2]. In obese patients who also have T2D, more than three-fourths of patients show remission after bariatric surgery [3]. Although remission rates may differ between different surgical procedures, high remission rates have been reported for Roux-en-Y gastric bypass [1, 3]. While many patients experience remission of diabetes, age, as well as duration and severity of disease have been presented as factors associated with reduced chance of achieving remission [1, 4]. Prediction of diabetes remission can be helpful in the clinical preoperative consultation and decision-making, and several indices have been constructed for this purpose. Scores like DiaRem [5], Ad-DiaRem [6], DiaBetter [7], the individualized metabolic surgery (IMS) score [8], and the age, body mass index, C-peptide level, and duration of T2D (ABCD) score [9] have been used for predicting diabetes remission after bariatric surgery. Many of the models based on the scores have high predictive capability and may already provide clinical guidance [10]. These tools might be helpful for personalized management of morbidly obese individuals with diabetes when considering bariatric surgery in routine care, ultimately contributing to precision medicine [11]. However, the performance of the scores in various studies was not consistent [6]. Previous prediction models were either limited by the small sample sizes or not validated using external data that were not seen by the models during model construction. Therefore, both the performance and validity of the models or scores are needed to be further evaluated and improved using larger database of bariatric surgery. In recent years, there have been a number of attempts to use artificial intelligence (AI) algorithms, including support vector machine (SVM) [12], decision tree [13], random forest [14], and deep learning algorithms such as artificial neural networks [15, 16] to incorporate preoperative predictors to predict outcomes of bariatric surgery. Compared with the traditional statistical regression models, AI algorithms have shown great promise in the field of bariatric surgery [17, 18]. However, to our knowledge, none have so far reached clinical practice. The aim of the present study was to construct a prediction model for diabetes remission using a deep learning AI algorithm, i.e. convolution neural network (CNN), and to compare its predictive capability comparing with four widely used predictive scores.

## 2. Methods

### 2.1. Study subjects

The study used the data from the Scandinavian Obesity Surgery Register (SOReg), a validated, national quality register covering virtually all bariatric and metabolic surgical procedures in Sweden [19]. By using the unique Swedish personal identification number, we linked SOReg to the Swedish National Patient Register, the Swedish National Death Register, the Swedish Prescribed Drug Register, and the Statistics Sweden to obtain information on inpatient and outpatient hospital visits, mortality, dispensed drugs, and individual socioeconomic data. The inclusion criteria for patients registered in SOReg were (1) operated with a primary Roux-en-Y gastric bypass (RYGB) procedure; and (2) diagnosed with T2D preoperatively, as defined by the American Diabetes Association [i.e. fasting plasma glucose ≥ 126 mg/L (7.0 mmol/L), Hemoglobin A1c (HbA1c) ≥ 48 mmol/mol (6.5%), or pharmacological treatment for diabetes] [20].

### 2.2. Outcome and predictor variables

The main outcome measure was complete remission of diabetes two years after surgery, defined as being without diabetes medication within a time frame of ± 6 months, i.e. 18 - 30 months postoperatively with normal HbA1c value <42 mmol/mol (6.0%) in accordance with the definitions of the American Diabetes Association [21]. Due to loss of information of HbA1c at follow-up, analyses of a secondary outcome, complete remission, defined as discontinuance of pharmacological treatment from 18-30 months was performed.

The predictor variables were patients’ demographic and socioeconomic information including age, sex, education level (primary, secondary, higher education < 3years, and high education ≥ 3 years), and region of residence characteristics (large city, medium city/town, and small town or rural area); preoperative body mass index (BMI), HbA1c, and treatment information including insulin treatment, metformin use, other non-insulin pharmacological treatment, and number of antidiabetic drugs; and preoperative comorbidities including sleep apnea, hypertension, dyslipidemia, depression, and cardiovascular comorbidity.

### 2.3. Descriptive analysis

Continuous variables were presented as mean ± standard deviation (SD), and the ordered and nominal variables were presented as median and interquartile range (IQR) and count and percentage (%), respectively. For comparison between two groups, the Student’s t-test and Mann-Whitney U test were used for continuous and ordered variables, respectively; and the Pearson’s chi-squared test was used for categorical variables. A two-tailed p-values < 0.05 was considered as statistically significant.

### 2.4. Multiple imputation for missing values

Missing values were assumed missing at random (MAR) and imputed using a random forest algorithm, which has the desirable properties of being able to handle mixed types of missing data, adaptive to interactions and nonlinearity, and potential to scale to big data settings [22]. To allow for the uncertainty of the imputation, in total 100 imputed datasets were generated in the current study.

### 2.5. Data normalization

Because the range of values of variables, such as age and BMI, varies widely, in some machine learning (ML) algorithms, objective function will not work properly [23]. Therefore, the continuous and ordered variables were normalized to have a mean of 0 and a standardization of 1, and the multi-category nominal variables (education and residence) were converted into several binary variables before they entered the ML models [24].

### 2.6. Predictive model

In the current study, we used a 7-layer convolution neural network (CNN) model with two one-dimensional (1D) convolution layers (with 100 filters for each), two 1D max pooling layers, one flatten layer, and two dense layers (with 1000 computation units) [25, 26]. The rectified linear unit activation function was used for the two 1D convolution layers and the first dense layers, and the sigmoid activation function was used for the last dense layer. The binary cross-entropy loss function and the adaptive moment estimation (Adam) optimizer were used when compiling the model [27].

### 2.7. Model training, validation, and test

The whole dataset was randomly split into two parts: a training dataset with 80% of the patients and a test dataset with 20% of the patients. During the model training stage, the training dataset was further split two datasets: one dataset with 64% of the patients to train the CNN model, and another with 16% of the patients to validate the model. Finally, the model was tested using the test dataset that was never seen by the CNN model. The CNN model was trained, validated, and tested with the 100 imputations. (Figure 1).

**Figure 1.**
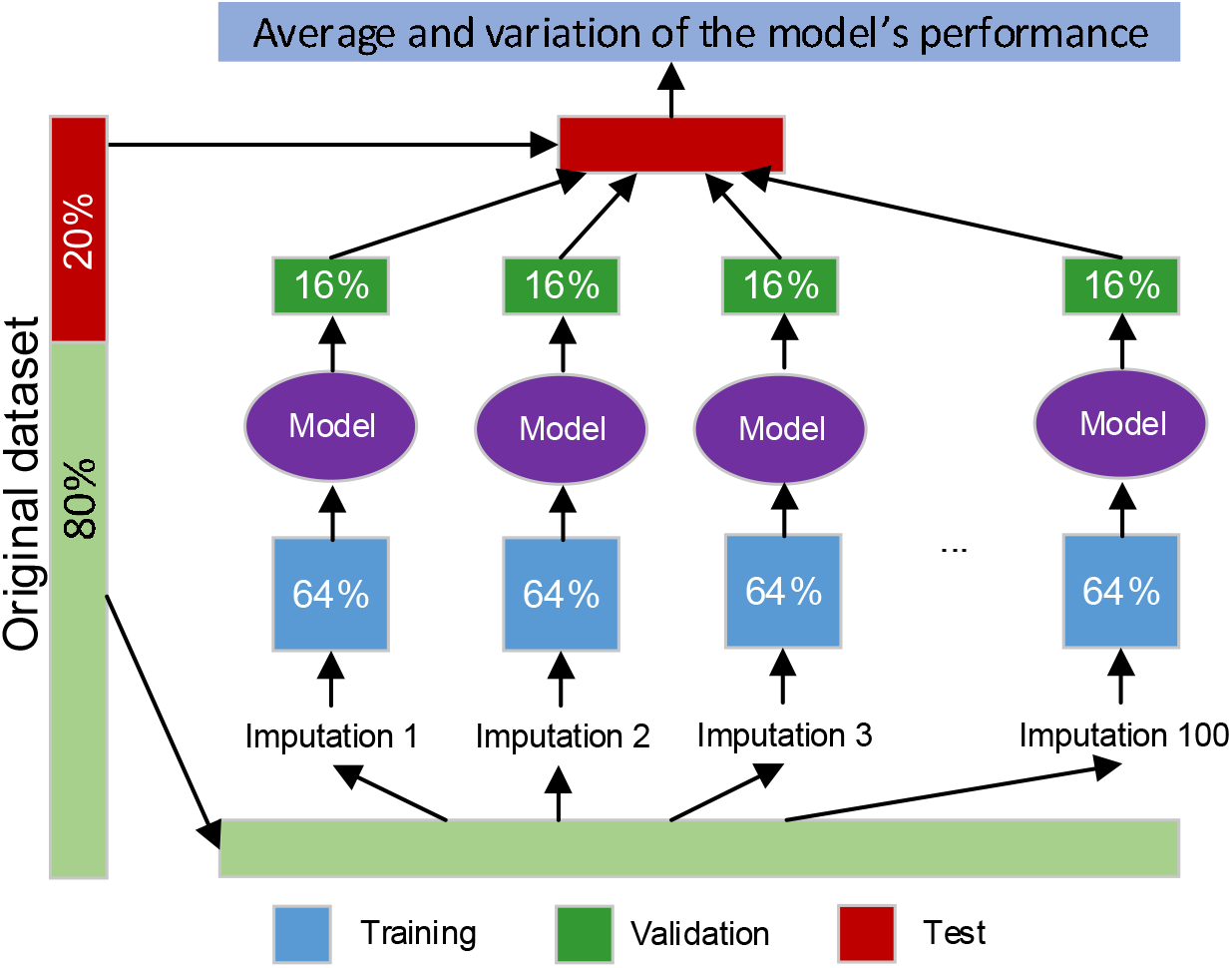
Procedure for training, validation, and testing for the CNN model

### 2.8. Indices of predictive ability

Predictive ability of the CNN model was evaluated using the following indices: area under the receiver operating characteristic (ROC) curve, sensitivity, specificity, and the Yunden J [28]. Terminology and derivations of the values were presented in detail previously [17]. The sensitivity and specificity presented in the study are the values on the ROC curve where the Youden J achieves the maximum value. The acceptable, excellent, and outstanding predictive models for were defined as the area under the ROC curve (AUC) of a model greater than 0.7, 0.8, and 0.9, respectively [29, 30]. The average and the 95% confidence interval (CI) of the indices were calculated based on the 100 imputations.

### 2.9. Comparison between the CNN model and DiaRem, Ad-DiaRem, DiaBetter, and IMS

We also evaluated the predictive capability of the currently used indices DiaRem, Ad-DiaRem, DiaBetter, and IMS, and compared them with the CNN model. The DiaRem score is calculated using insulin use, age, HbA1c value, and type of antidiabetic drugs [31]. The Ad-DiaRem score is a modification of the DiaRem score, calculated using insulin use, age, HbA1c value, number of antidiabetic drugs, duration of diabetes, and number of antidiabetic drugs [12]. The DiaBetter is calculated using HbA1c, type of antidiabetic drugs, and duration of diabetes [7]. The IMS score is calculated using the number of preoperative diabetes medications, insulin use, duration of diabetes, and HbA1c level [8].

The points on the nonparametric ROC curve of DiaRem, Ad-DiaRem, DiaBetter, and IMS were generated using each value as a classification cut-point and computing the corresponding sensitivity and one minus specificity. These points were then connected by straight lines, and the area under the resulting ROC curve was computed using the trapezoidal rule [32].

The same training and testing procedure used for the CNN model was applied for the four scores as well.

### 2.10. Software and hardware

The descriptive analysis and evaluation for DiaRem, Ad-DiaRem, DiaBetter, and IMS were conducted in Stata 16.1 (StataCorp LLC, College Station, TX, USA). The CNN model was achieved in Python 3.6 (Python Software Foundation, https://www.python.org/) using packages Keras 2.4.0 and Scikit-learn 0.23. All the computation was operated on a computer with 64-bit Windows 7 Enterprise operating system (Service Pack 1), Intel ® Core TM i5-4210U CPU of 2.40 GHz, and 16.0 GB installed random access memory.

### 2.11. Ethics

The study was approved by the regional ethics committee in Stockholm (reference numbers: 2013/535-31/5, 2014/1639-32, and 2017/857-32).

## 3. Results

### 3.1. Characteristics of the patients

In total, 8112 patients met the inclusion criteria, after exclusion of 55 patients who died during the first two years after surgery, 8057 patients remained in the current analysis. Information on pharmaceutical usage before and after surgery was available for all patients. A postoperative weight was registered for 7268 patients at one year after surgery (90.2%), and 4996 patients at two years after surgery (62.0%). A postoperative glycosylated HbA1c test result was available for 6989 patients (86.7%). Baseline characteristics of the included patients are shown in Table 1. Statistically significant differences were found for almost all the predictor variables between the remission patients and non-remission patients, except for depression and education (Table 1), which implies the potential of using the predictor variables to predict the outcome. Preoperative HbA1c-values were missing for about one seventh of the patients, indicating the need for imputation since the predictive capability otherwise would be significantly reduced and biased by excluded the considerable proportion of the data with missing values. Patients with a missing HbA1c value were more often males of marginally higher age and longer duration of disease, small differences were also seen in terms of pharmacological treatment, education and residence (Supplementary Table S1). After multiple imputation, similar distributions of HbA1c values were seen (Supplementary Figure S1).

**Table 1.**
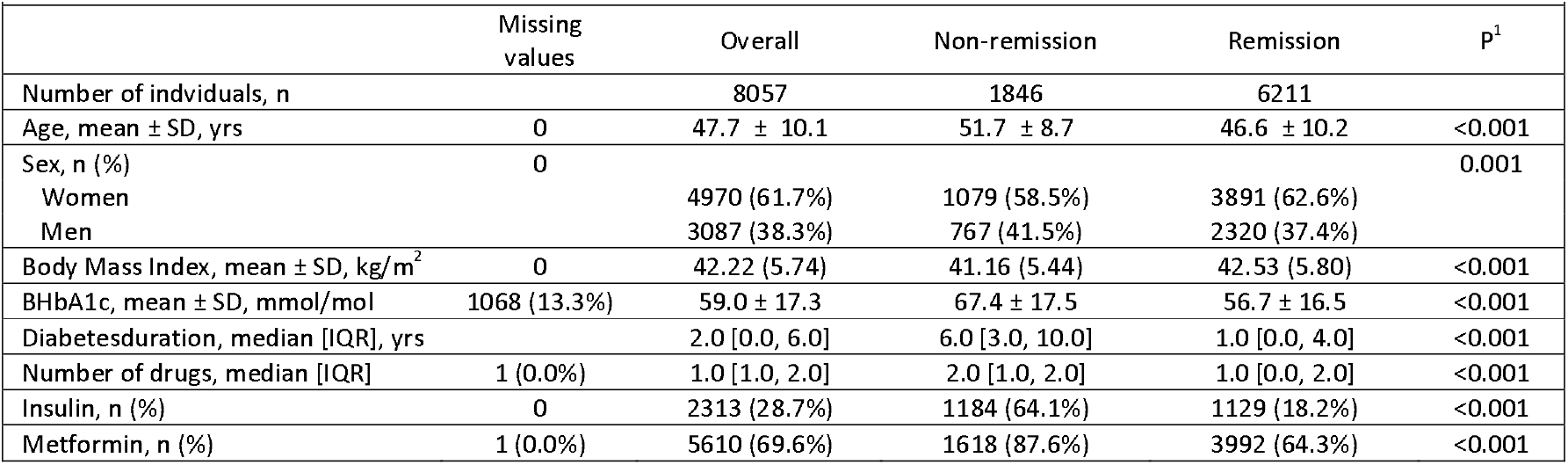
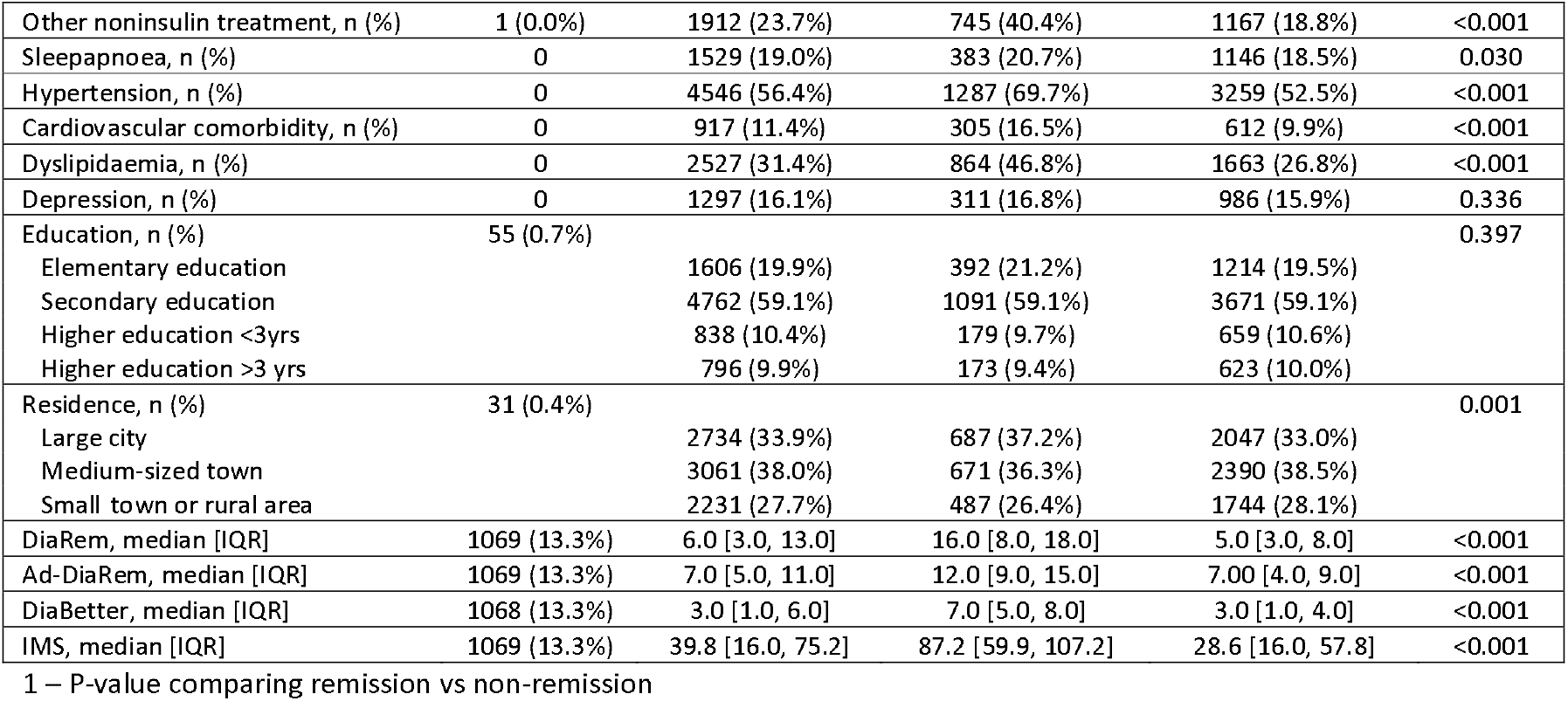
Characteristics of study participants with further stratification on remission of diabetes

### 3.2. Surgical outcome

Mean BMI-loss at 1 year after surgery was 12.2 ± 4.0 kg/m^2^, with excess BMI loss [EBMIL = 100 × (initial BMI – postoperative BMI)/(initial BMI – 25) %] of 74.0 ± 22.5%, and total weight loss (TWL = 100 × weight loss/preoperative weight %) of 28.7 ± 7.6%. Mean BMI-loss at two years after surgery was 12.0 ± 4.53 kg/m^2^, with EBMIL of 73.3 ± 24.4%, and TWL of 28.4 ± 8.9%. At 2 years after surgery 77.1% (n=6211) of the 8057 patients were able to discontinue pharmacological treatment at two years after surgery, while complete remission was seen for 62.2% (n=4004) of the 6348 patients who had been evaluated for complete remission.

### 3.3. Predictive capability of the CNN model, DiaRem, Ad-DiaRem, Diabetter, and IMS

The predictive capability of the CNN model for the major outcome (remission) is shown in Figure 2 and Table 2. In both the trainings and validations, the CNN model presented good predictive ability, with an AUC of 0.86 (95% CI: 0.85, 0.87) and 0.85 (95% CI: 0.83, 0.86), respectively (Table 2).

**Table 2.**
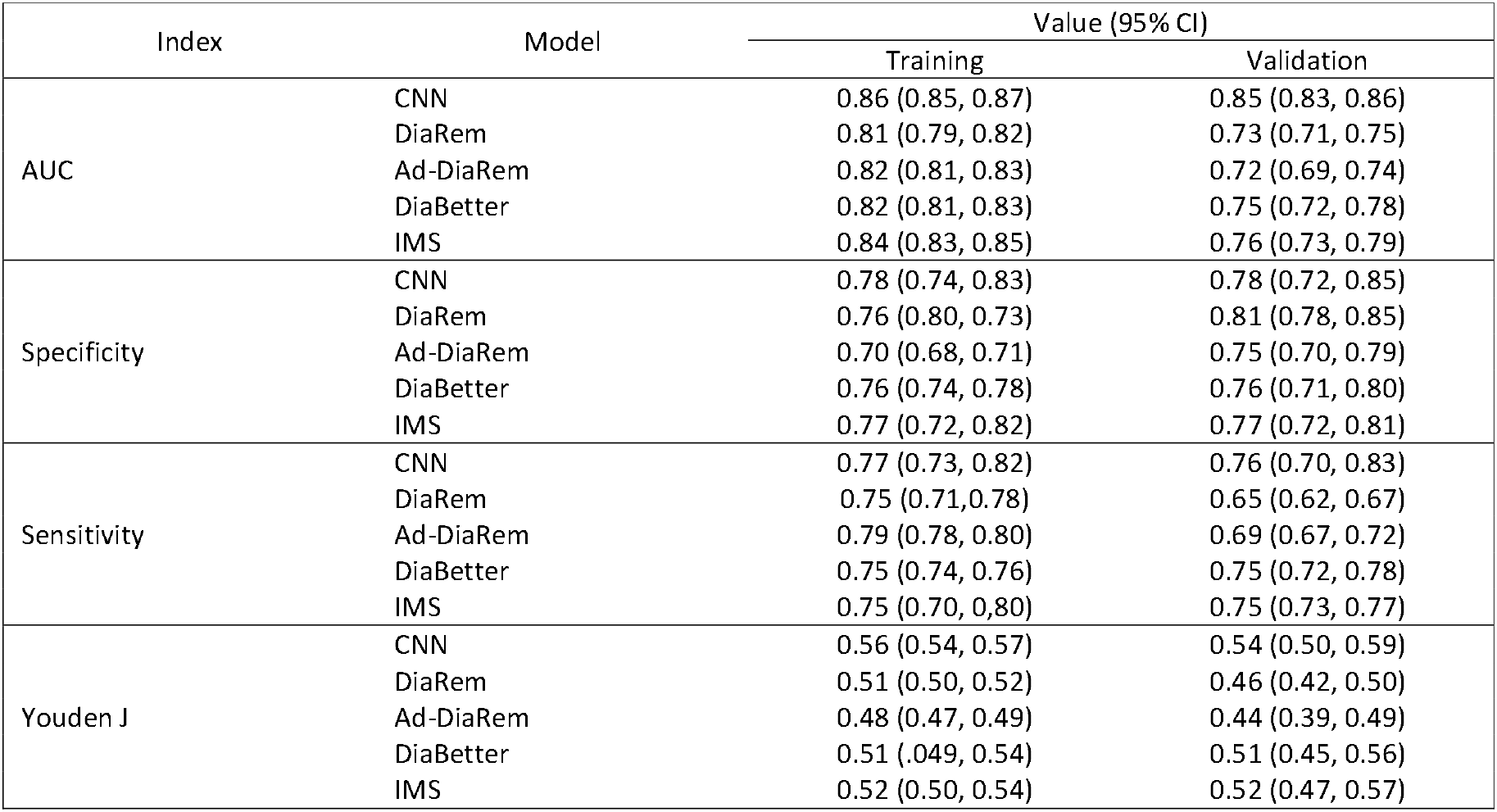
Predictive capability of the CNN model and diabetes indices for the major outcome

|  |  |  |  |
| --- | --- | --- | --- |
| Index | Model | Value (95% CI) |  |
|  |  | Training | Validation |
| AUC | CNN | 0.86 (0.85, 0.87) | 0.85 (0.83, 0.86) |
|  | DiaRem | 0.81 (0.79, 0.82) | 0.73 (0.71, 0.75) |
|  | Ad-DiaRem | 0.82 (0.81, 0.83) | 0.72 (0.69, 0.74) |
|  | DiaBetter | 0.82 (0.81, 0.83) | 0.75 (0.72, 0.78) |
|  | IMS | 0.84 (0.83, 0.85) | 0.76 (0.73, 0.79) |
| Specificity | CNN | 0.78 (0.74, 0.83) | 0.78 (0.72, 0.85) |
|  | DiaRem | 0.76 (0.80, 0.73) | 0.81 (0.78, 0.85) |
|  | Ad-DiaRem | 0.70 (0.68, 0.71) | 0.75 (0.70, 0.79) |
|  | DiaBetter | 0.76 (0.74, 0.78) | 0.76 (0.71, 0.80) |
|  | IMS | 0.77 (0.72, 0.82) | 0.77 (0.72, 0.81) |
| Sensitivity | CNN | 0.77 (0.73, 0.82) | 0.76 (0.70, 0.83) |
|  | DiaRem | 0.75 (0.71, 0.78) | 0.65 (0.62, 0.67) |
|  | Ad-DiaRem | 0.79 (0.78, 0.80) | 0.69 (0.67, 0.72) |
|  | DiaBetter | 0.75 (0.74, 0.76) | 0.75 (0.72, 0.78) |
|  | IMS | 0.75 (0.70, 0.80) | 0.75 (0.73, 0.77) |
| Youden J | CNN | 0.56 (0.54, 0.57) | 0.54 (0.50, 0.59) |
|  | DiaRem | 0.51 (0.50, 0.52) | 0.46 (0.42, 0.50) |
|  | Ad-DiaRem | 0.48 (0.47, 0.49) | 0.44 (0.39, 0.49) |
|  | DiaBetter | 0.51 (0.49, 0.54) | 0.51 (0.45, 0.56) |
|  | IMS | 0.52 (0.50, 0.54) | 0.52 (0.47, 0.57) |

**Figure 2.**
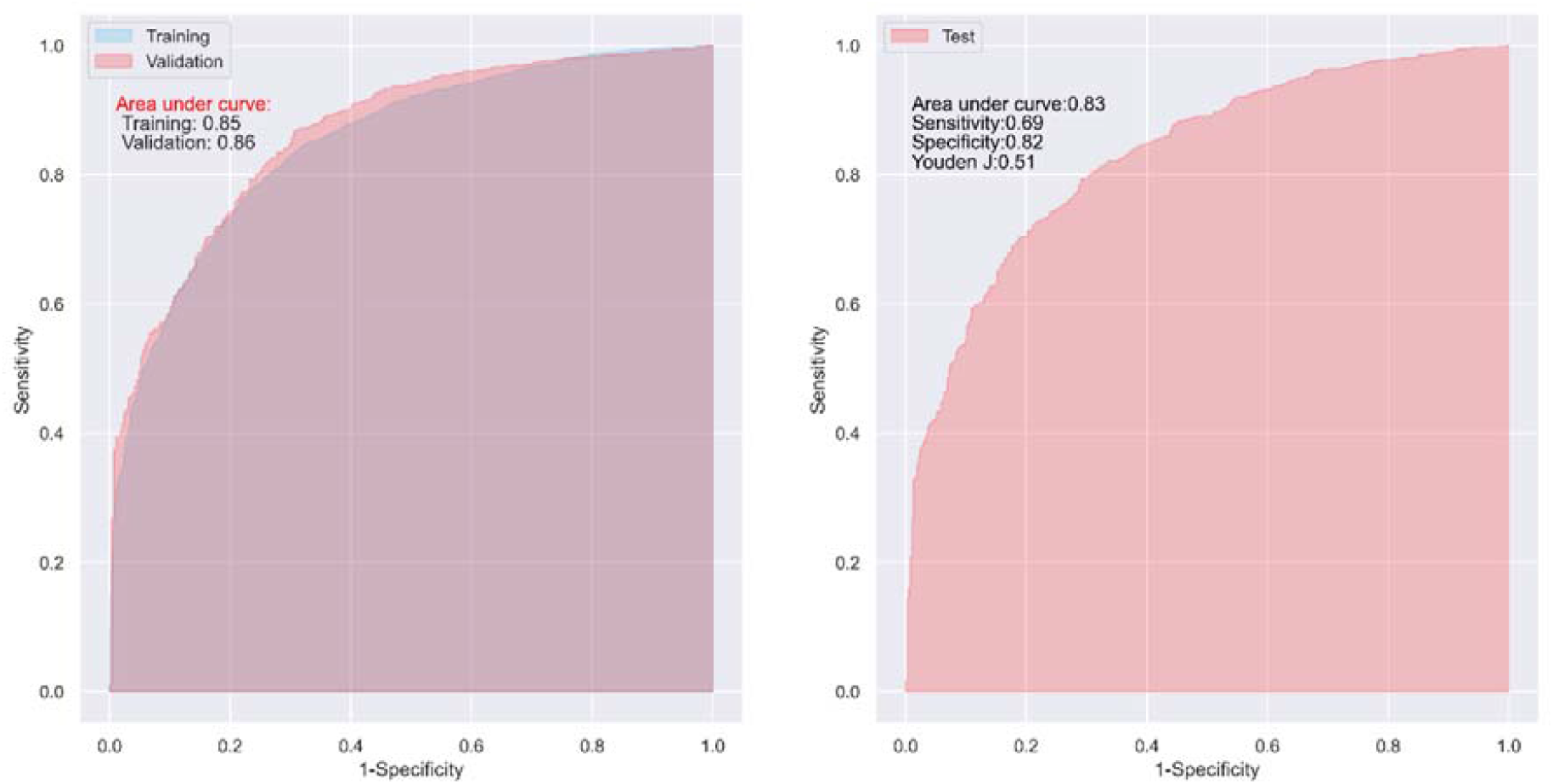
ROC curves of the CNN model in one of the 100 trainings and validations (left), and tests (right)

The DiaRem, Ad-DiaRem, DiaBetter, and IMS also showed good predictive capability in the training with an AUC > 0.8 (Figure 3 left and Table 2), but only acceptable predictive ability in the validations (Table 2), with an AUC of 0.73 (95% CI: 0.71, 0.75), 0.72 (95% CI: 0.69, 0.74), 0.75 (95% CI: 0.72, 0.78), and 0.76 (95% CI: 0.73, 0.79), respectively. In general, the predictive capability of the CNN model was 16.4%, 18.1%, 13.3%, and 11.8% higher than that of DiaRem, Ad-DiaRem, DiaBetter, and IMS, in terms of AUC, respectively. In the tests, the predictive ability of the CNN model was 10.6%, 12.2%, 12.2%, and 9.2% higher than that of DiaRem, Ad-DiaRem, DiaBetter, and IMS in terms of AUC [AUC = 0.83 (95% CI: 0.82, 0.85) vs. 0.75 (95% CI: 0.73, 0.76), 0.74 (95% CI: 0.71, 0.77), 0.74 (95% CI: 0.72, 0.76), and 0.76 (95% CI: 0.73, 0.78)], respectively (Figure 2 right and Figure 3 right).

**Figure 3.**
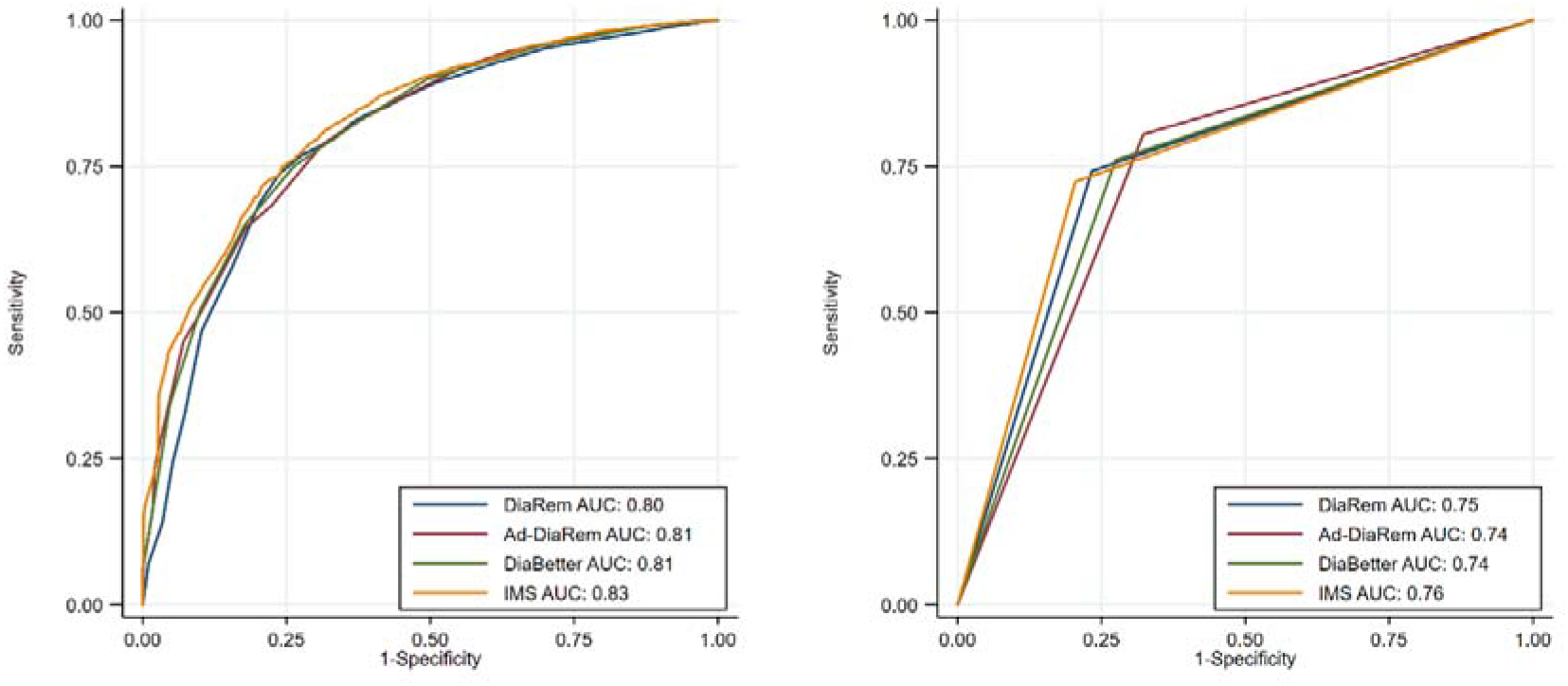
ROC curves of diabetes indices in one of the 100 trainings (left), and tests (right)

For the secondary outcome, complete remission, the CNN model also presented a good predictive capability in both the trainings and validations, with an AUC of 0.84 (95% CI: 0.83, 0.85) and 0.83 (95% CI: 0.81, 0.85), respectively (Supplementary Table S4). Although DiaRem, Ad-DiaRem, DiBbetter, and IMS showed good predictive ability in the training with an AUC ≥0.80, they only showed acceptable predictive ability in the validations with an AUC of 0.72 (95% CI: 0.69, 0.75), 0.72 (95% CI: 0.69, 0.74), 0.74 (95% CI: 0.72, 0.77), and 0.74 (95% CI: 0.72, 0.76), respectively (Supplementary Table S4). In general, the predictive capability of the CNN model was 15.3%, 15.3%, 12.2%, and 12.2% higher than that of DiaRem, Ad-DiaRem, DiaBetter, and IMS, in terms of AUC, respectively.

In the tests, the predictive capability of the CNN model was 9.3%, 10.8%, 10.8%, and 9.3% higher than that of DiaRem, Ad-DiaRem, DiaBetter, and IMS in terms of AUC [AUC = 0.82 (95% CI: 0.81, 0.83) vs. 0.75 (95% CI: 0.73, 0.78), 0.74 (95% CI: 0.73, 0.75), 0.74 (95% CI: 0.71, 0.76), and 0.75 (95% CI: 0.73, 0.77)], respectively (Supplementary Figure S2 right and S3 right).

## 4. Discussion

The preoperative decision process before bariatric surgery is highly complex and involves issues including perioperative safety, long-term complications as well as efficacy in terms of weight-loss and improvement of obesity-related comorbidities, but also concerns and fears of the patient. Using preoperative information to predict diabetes remission after bariatric surgery may help clinicians make an optimal decision for diabetes treatment by balancing the surgical risks against the potential benefit. An ideal model should be sensitive enough to discriminate the candidate T2D patients who might benefit from the surgery, and should be consistent and reproducible in patients with different characteristics [33]. Previous studies have tried to identify preoperative predictors of diabetes remission following bariatric surgery in T2D patients, and some common factors emerged, including age, BMI; duration of diabetes, HbA1c level, and insulin use [12, 13, 31, 34-39].

The currently available and widely accepted predictive indices for diabetes remission, including DiaRem, Ad-DiaRem, DiaBetter, and IMS used in our study, are all simple and easily available to clinicians as clinical decision support. In addition, one other index, the ABCD score [40], also included c-peptide. This laboratory measure would add information of endogenous insulin production and could thus potentially further enhance the effectiveness of a prediction model. However, the ABCD score has not been shown to have higher predictive capacity to other available models, and it is highly possible that other measures of severity of T2D disease, such as duration of disease, HbA1c-value, and type and number of drugs, may provide the same or even better measures for a prediction model [10]. The performance of the comprehensive indices varies in the previous studies, with the AUC ranging from 0.70 to 0.95, due to the differences in remission rate (ranging from 24% to 84%), type of surgery, baseline characteristics of the patients, follow-up duration, and even the definition of remission [5, 41-44]. Besides the aforementioned heterogeneities, the previous studies also suffered the limitation of small sample size; most of them only included one or two hundred patients, and few had more than 500 patients.

The use of AI or machine learning techniques in medical research and practice is currently an evolving topic with great potential. While the exact role of AI in this setting remains to be established, one potential area where AI seems to outperform traditional techniques is in the construction of prediction models for outcomes from surgical procedures [45]. Previous studies have reported promising results using sparse SVM, decision tree, and artificial neural network for prediction of diabetes remission after bariatric surgery [12],[13],[16]. However, all of these studies also have the same limitation, i.e. small sample size (ranging from 130 to 352), therefore the external validity of the models is questionable.

To our knowledge, our study is the largest that included more than 8,000 T2D patients in the national register SOReg to explore the potential of the AI algorithm CNN in predicting diabetes remission after bariatric surgery. The CNN model showed higher accuracy for cessation of antidiabetic drugs as well as for complete remission in both training, validation and the final test. While the four established predictive indices based on traditional statistic methods also performed well, the CNN model outperformed the four indices by around 10% in the final test. The large dataset and robust results indicate that the model may constitute a valuable tool in the preoperative setting, taking advantage of the AI algorithm, and may behave better if being trained using more patients and/or preoperative variables.

Some of the postoperative outcomes of bariatric surgery involves several factors that cannot be estimated preoperatively. Previous construction of prediction models for perioperative complications have reported discouraging results, mainly as a direct cause of the complexity and diversity of causes for perioperative complications [17, 26, 46]. In contrast with safety outcomes, efficacy outcomes (in particular those of highly standardized surgical methods such as gastric bypass) may be more suited for adequate prediction models since the factors influencing long-term effects are less diverse. Remission of diabetes is one such outcome that to large extent is influenced by a few specific factors, making prediction models more easily available.

Our CNN model included measures of patient-specific characteristics, information on duration and severity of disease, as well as a few socioeconomic factors that all should be easily available at the time of consultation before surgery. While it is likely that the model could have reached a higher precision if postoperative results (such as early weight-loss or improvement in glucose homeostasis) were included, these measures are not available in the preoperative setting and their inclusion would therefor reduce the clinical usefulness of the model [1, 4, 47].

The CNN model or other neural networks have a, although finite, capacity (depending on their storage) to remember what they learned before. For example, we conducted an altered model training strategy. Once the model was constructed, we trained, validated and tested it sequentially using randomly selected patients from the dataset (Supplementary Figure S4).

We can imagine that with the increased training numbers, increasingly more or even all the data would be seen by the model, and old parameters would be updated while new parameters would be generated, and therefore its predictive ability would increase gradually. In our model, we observed that with the increased training number, sensitivity, specificity, AUC, and Youden J of the model all approached 1 in the final test (Supplementary Figure S5 left). However, this capacity does not exist in traditional regression models, such as logistic regression, where the predictive ability keeps constant regardless of the training number (Supplementary Figure S5 right). This would imply that with more available data would improve the predictive capability of the CNN model even further, at least in the Swedish context. To generalize the application of the CNN model, a multinational registration consortium of gastric bypass surgery patients would be needed for the improved model training, but also for validation.

However, the capacity of memory is also a limitation of the CNN, because it reduces the model’s flexibility to incorporate the information from external unseen data, and therefore results in overfitting to specific past data, underfitting to the new data, and impedes generalization of the model [48]. Teaching neural networks to forget strategically is an important task in ML. This highlights one of the major challenge of machine learning techniques [49]. To fulfill this task, incorporating long short-term memory units into CNN networks has been attempted to process temporal sequences and reduce model parameters in human face and activity recognition, which has shown consistent superior performance and good generalization [50, 51]. Furthermore, The transparency of ML methods is more complex than that of traditional regression models, which makes them harder for researchers to understand the exact model reported [48]. In the absence of clear guidelines, we have – to the best of our ability-conducted and reported the study to match the requirements of the TRIPOD statement as

well as suggested modifications [52, 53]. The programming code of the study is available at the repository website figshare [54]. In addition, the study was only based on data from one nation. For full use of the model, external validation would be needed in other parts of the world as well. Despite these potential limitations, the CNN model outperformed the currently available high-quality prediction models. It also reported better predictive ability to that of a previous report on AI for diabetes remission [55]. The CNN model may therefor find a place in the preoperative setting for surgeons, bariatricians, or endocrinologists looking to quantify the chance of diabetes remission in their decision for bariatric surgery for a specific patient.

## 5. Conclusion

Our CNN based ML model performed well in identifying morbidly obese patients with T2D who might benefit from Roux-en-Y gastric bypass surgery. We also demonstrated the model had better predictive capability compared with the currently widely used four comprehensive indices for diabetes remission after gastric bypass surgery. Prospectively identifying this subset of patients using data available at the time of preoperative evaluation provides an opportune time window to intervene and prevent or reduce the risk of morbidity and mortality, and potentially reduce total cost of care. However, this model should be further validated in future research using external data in other countries before incorporating in clinical practice.

## Data Availability

The data that support the study are not publicly available because they contain information that could compromise research participant privacy and confidentiality. The authors will make the data available upon reasonable request and with permission of the Committee of Scandinavian Obesity Surgery Registry in Orebro, Sweden.

## Data Availability Statement

The data that support the study are not publicly available because they contain information that could compromise research participant privacy and confidentiality. The authors will make the data available upon reasonable request and with permission of the Committee of Scandinavian Obesity Surgery Registry in Örebro, Sweden.

## Supplemental materials

**Supplementary Table S1.** Characteristics of study participants stratification on availability of preoperative HbA1c analysis.

|  |  |  |  |
| --- | --- | --- | --- |
|  | Available HbA1c | Non-available HbA1c | p |
| Number of individuals, n | 6989 | 1068 |  |
| Age, mean ± SD, yrs | 47.7 ± 10.1 | 48.0 ± 10.0 | 0.428 |
| Sex, n (%) |  |  |  |
| Women | 4343 (62.1) | 627 (58.7) | 0.034 |
| Men | 2646 (37.9) | 441 (41.3) |  |
| Body Mass Index, mean ± SD, kg/m<sup>2</sup> | 42.20 ± 5.74 | 42.31 ± 5.75 | 0.576 |
| Diabetes duration, median [IQR], yrs | 2.0 [0.0, 6.0] | 3.0 [1.0, 6.0] | <0.001 |
| Number of drugs, median [IQR] | 1.0 [1.0, 2.0] | 1.0 [1.0, 2.0] | <0.001 |
| Insulin, n (%) | 1915 (27.4) | 398 (37.3) | <0.001 |
| Metformin, n (%) | 4740 (67.8) | 870 (81.5) | <0.001 |
| Other noninsulin treatment, n (%) | 1597 (22.9) | 315 (29.5) | <0.001 |
| Sleepapnoea, n (%) | 1323 (18.9) | 206 (19.3) | 0.813 |
| Hypertension, n (%) | 3963 (56.7) | 583 (54.6) | 0.206 |
| Cardiovascular comorbidity, n (%) | 779 (11.1) | 138 (12.9) | 0.099 |
| Dyslipidaemia, n (%) | 2196 (31.4) | 331 (31.0) | 0.806 |
| Depression, n (%) | 1132 (16.2) | 165 (15.4) | 0.566 |
| Education, n (%) |  |  | 0.034 |
| Elementary education | 4090 (58.5) | 672 (62.9) |  |
| Secondary education | 1398 (20.0) | 208 (19.5) |  |
| Higher education <3yrs | 744 (10.6) | 94 (8.8) |  |
| Higher education >3 yrs | 710 (10.2) | 86 (8.1) |  |
| Residence, n (%) |  |  | <0.001 |
| Large city | 2481 (35.5) | 253 (23.7) |  |
| Medium-sized town | 2718 (38.9) | 343 (32.1) |  |
| Small town or rural area | 1762 (25.2) | 469 (43.9) |  |

**Supplementary Table S2.** Characteristics of the patients having complete remission information vs. without complete remission information.

|  |  |  |  |
| --- | --- | --- | --- |
| Variable | Information available | Information missing | p |
| Number of individuals, n | 6438 | 1674 |  |
| Age, mean ± SD, yrs | 48.3 ± 10.0 | 46.0 ± 10.4 | <0.001 |
| Sex, n (%) |  |  | 0.002 |
| Women | 4022 (62.5) | 976 (58.3) |  |
| Men | 2416 (37.5) | 698 (41.7) |  |
| Body Mass Index, mean ± SD, kg/m<sup>2</sup> | 42.2 ± 5.7 | 42.3 ± 5.8 | 0.622 |
| B-HbA1c mean ± SD, mmol/mol | 59.00 ± 17.3 | 59.2 ± 17.6 | 0.752 |
| Diabetesduration, median [IQR], yrs | 2.0 [0.0, 6.0] | 2.0 [0.0, 5.0] | <0.001 |
| Number of drugs, median [IQR] | 1.0 [1.0, 2.0] | 1.00 [1.0, 2.0] | 0.048 |
| Insulin, n (%) | 1861 (28.9) | 469 (28.0) | 0.492 |
| Metformin, n (%) | 4501 (69.9) | 1147 (68.5) | 0.472 |
| Other noninsulin treatment, n (%) | 1559 (24.2) | 363 (21.7) | 0.083 |
| Sleepapnoea, n (%) | 1240 (19.3) | 307 (18.3) | 0.412 |
| Hypertension, n (%) | 3742 (58.1) | 841 (50.2) | <0.001 |
| Cardiovascular comorbidity, n (%) | 746 (11.6) | 190 (11.4) | 0.820 |
| Dyslipidaemia, n (%) | 2090 (32.5) | 461 (27.5) | <0.001 |
| Depression, n (%) | 982 (15.3) | 330 (19.7) | <0.001 |
| Education, n (%) |  |  | 0.002 |
| Elementary education | 3816 (59.3) | 969 (57.9) |  |
| Secondary education | 1244 (19.3) | 381 (22.8) |  |
| Higher education <3yrs | 692 (10.7) | 152 (9.1) |  |
| Higher education >3 yrs | 647 (10.0) | 154 (9.2) |  |
| Residence, n (%) |  |  | <0.001 |
| Large city | 2242 (34.8) | 504 (30.1) |  |
| Medium-sized town | 2411 (37.4) | 666 (39.8) |  |
| Small | 1764 (27.4) | 480 (28.7) |  |
| DiaRem, median [IQR] | 6.0 [3.0, 14.0] | 5.0 [3.0, 10.0] | <0.001 |
| Ad-DiaRem, median [IQR] | 7.0 [5.0, 11.0] | 7.0 [4.0, 10.0] | <0.001 |
| DiaBetter, median [IQR] | 3.0 [1.0, 6.0] | 3.0 [1.0, 5.0] | <0.001 |
| IMS, median [IQR] | 39.8 [16.0, 76.0] | 34.2 [16.0, 64.8] | <0.001 |

**Supplementary Table S3.** Characteristics of the patients complete remission vs. non complete remission.

|  |  |  |  |
| --- | --- | --- | --- |
| Variable | No | Yes | p |
| Number of individuals, n | 2434 | 4004 |  |
| Age, mean ± SD, yrs | 51.42 (8.59) | 46.32 (10.22) | <0.001 |
| Sex, n (%) |  |  | 0.136 |
| Women | 1492 (61.3) | 2530 (63.2) |  |
| Men | 942 (38.7) | 1474 (36.8) |  |
| Body Mass Index, mean ± SD, kg/m<sup>2</sup> | 41.16 (5.32) | 42.85 (5.88) | <0.001 |
| B-HbA1c mean ± SD, mmol/mol | 66.07 (17.37) | 54.85 (15.83) | <0.001 |
| Diabetes duration, median [IQR], yrs | 5.0 [2.0, 9.0] | 1.0 [0.0, 4.0] | <0.001 |
| Number of drugs, median [IQR] | 2.0 [1.0, 2.0] | 1.0 [0.0, 1.0] | <0.001 |
| Insulin, n (%) | 1306 (53.7) | 555 (13.9) | <0.001 |
| Metformin, n (%) | 1991 (81.8) | 2510 (62.7) | <0.001 |
| Other noninsulin treatment, n (%) | 936 (38.5) | 623 (15.6) | <0.001 |
| Sleepapnoea, n (%) | 494 (20.3) | 746 (18.6) | 0.107 |
| Hypertension, n (%) | 1657 (68.1) | 2085 (52.1) | <0.001 |
| Cardiovascular comorbidity, n (%) | 369 (15.2) | 377 (9.4) | <0.001 |
| Dyslipidaemia, n (%) | 1090 (44.8) | 1000 (25.0) | <0.001 |
| Depression, n (%) | 397 (16.3) | 585 (14.6) | 0.071 |
| Education, n (%) |  |  | 0.037 |
| Elementary education | 1453 (59.7) | 2363 (59.0) |  |
| Secondary education | 503 (20.7) | 741 (18.5) |  |
| Higher education <3yrs | 233 (9.6) | 459 (11.5) |  |
| Higher education >3 yrs | 230 (9.4) | 417 (10.4) |  |
| Residence, n (%) |  |  | 0.268 |
| Large city | 883 (36.3) | 1359 (33.9) |  |
| Medium-sized town | 885 (36.4) | 1526 (38.1) |  |
| Small | 659 (27.1) | 1105 (27.6) |  |
| DiaRem, median [IQR] | 14.0 [6.0, 17.0] | 4.0 [2.0, 7.0] | <0.001 |
| Ad-DiaRem, median [IQR] | 11.0 [8.0, 14.0] | 6.0 [3.0, 9.0] | <0.001 |
| DiaBetter, median [IQR] | 6.0 [4.0, 8.0] | 2.0 [1.0, 4.0] | <0.001 |
| IMS, median [IQR] | 77.4 [45.4, 102.6] | 27.2 [12.6, 48.6] | <0.001 |

**Supplementary Table S4.** Predictive capability of the CNN model and diabetes indices for complete remission.

|  |  |  |  |
| --- | --- | --- | --- |
| Index | Model | Value (95% CI) |  |
|  |  | Training | Validation |
| AUC | CNN | 0.84 (0.83, 0.85) | 0.83 (0.81, 0.85) |
|  | DiaRem | 0.80 (0.79, 0.81) | 0.72 (0.69, 0.75) |
|  | Ad-DiaRem | 0.81 (0.80, 0.82) | 0.72 (0.69, 0.74) |
|  | DiaBetter | 0.80 (0.79, 0.81) | 0.74 (0.72, 0.77) |
|  | IMS | 0.81 (0.79, 0.82) | 0.74 (0.72, 0.76) |
| Specificity | CNN | 0.76 (0.71, 0.80) | 0.76 (0.68, 0.83) |
|  | DiaRem | 0.70 (0.65, 0.75) | 0.72 (0.68, 0.76) |
|  | Ad-DiaRem | 0.71 (0.59, 0.84) | 0.75 (0.70, 0.79) |
|  | DiaBetter | 0.67 (0.66, 0.69) | 0.68 (0.64, 0.71) |
|  | IMS | 0.72 (0.68, 0.76) | 0.72 (0.68, 0.76) |
| Sensitivity | CNN | 0.78 (0.74, 0.83) | 0.77 (0.70, 0.84) |
|  | DiaRem | 0.77 (0.73, 0.80) | 0.73 (0.69, 0.76) |
|  | Ad-DiaRem | 0.75 (0.63, 0.87) | 0.69 (0.67, 0.72) |
|  | DiaBetter | 0.81 (0.80, 0.82) | 0.81 (0.78, 0.84) |
|  | IMS | 0.77 (0.73, 0.81) | 0.77 (0.74, 0.80) |
| Youden J | CNN | 0.54 (0.53, 0.55) | 0.53 (0.49, 0.57) |
|  | DiaRem | 0.47 (0.45, 0.49) | 0.44 (0.39, 0.50) |
|  | Ad-DiaRem | 0.46 (0.45, 0.48) | 0.44 (0.39, 0.49) |
|  | DiaBetter | 0.48 (0.46, 0.50) | 0.48 (0.43, 0.53) |
|  | IMS | 0.49 (0.47, 0.50) | 0.48 (0.43, 0.53) |

**Supplementary Figure S1.**
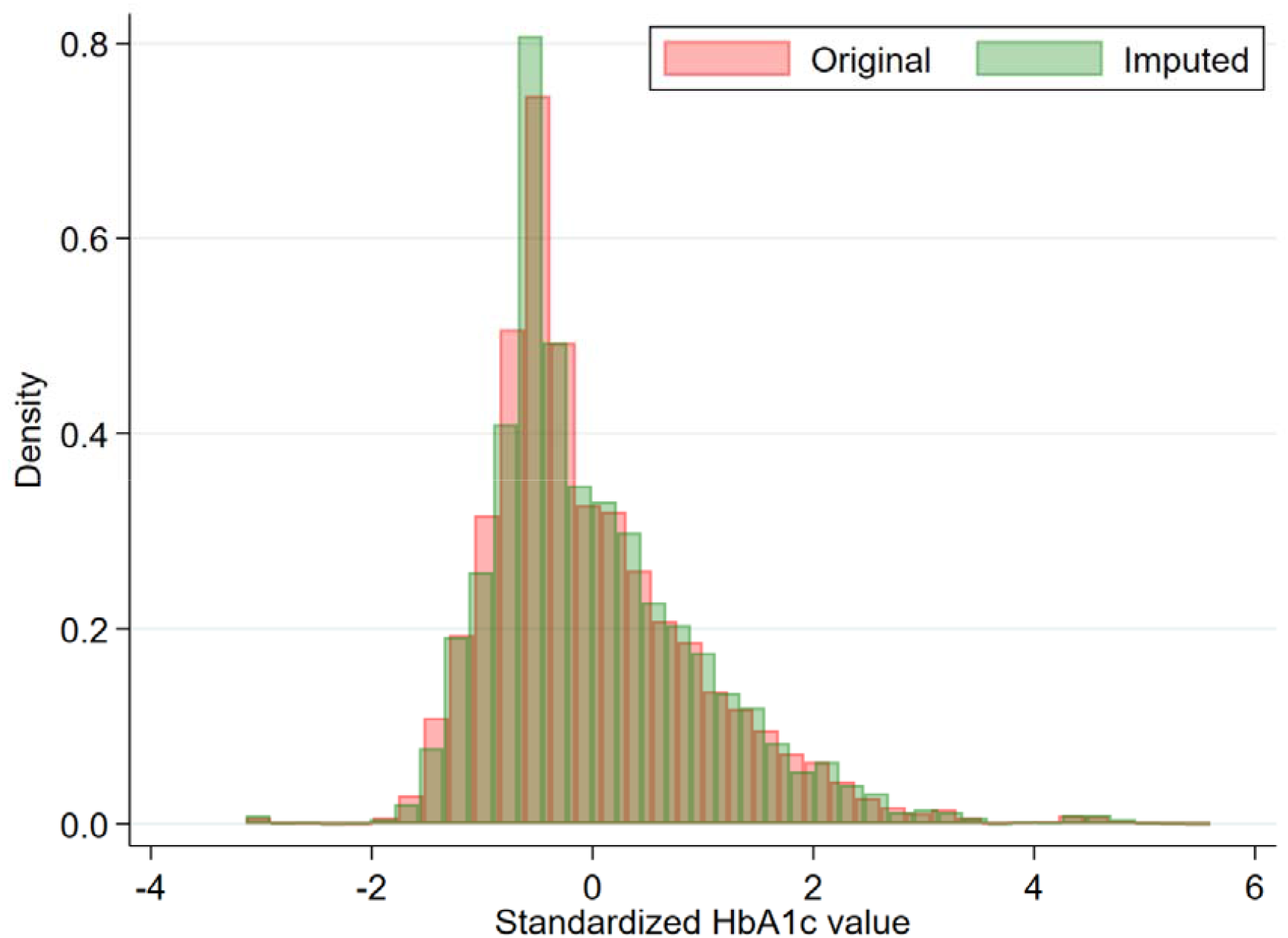
Distributions of the original HbA1c values and the HbA1c values after imputation

**Supplementary Figure S2.**
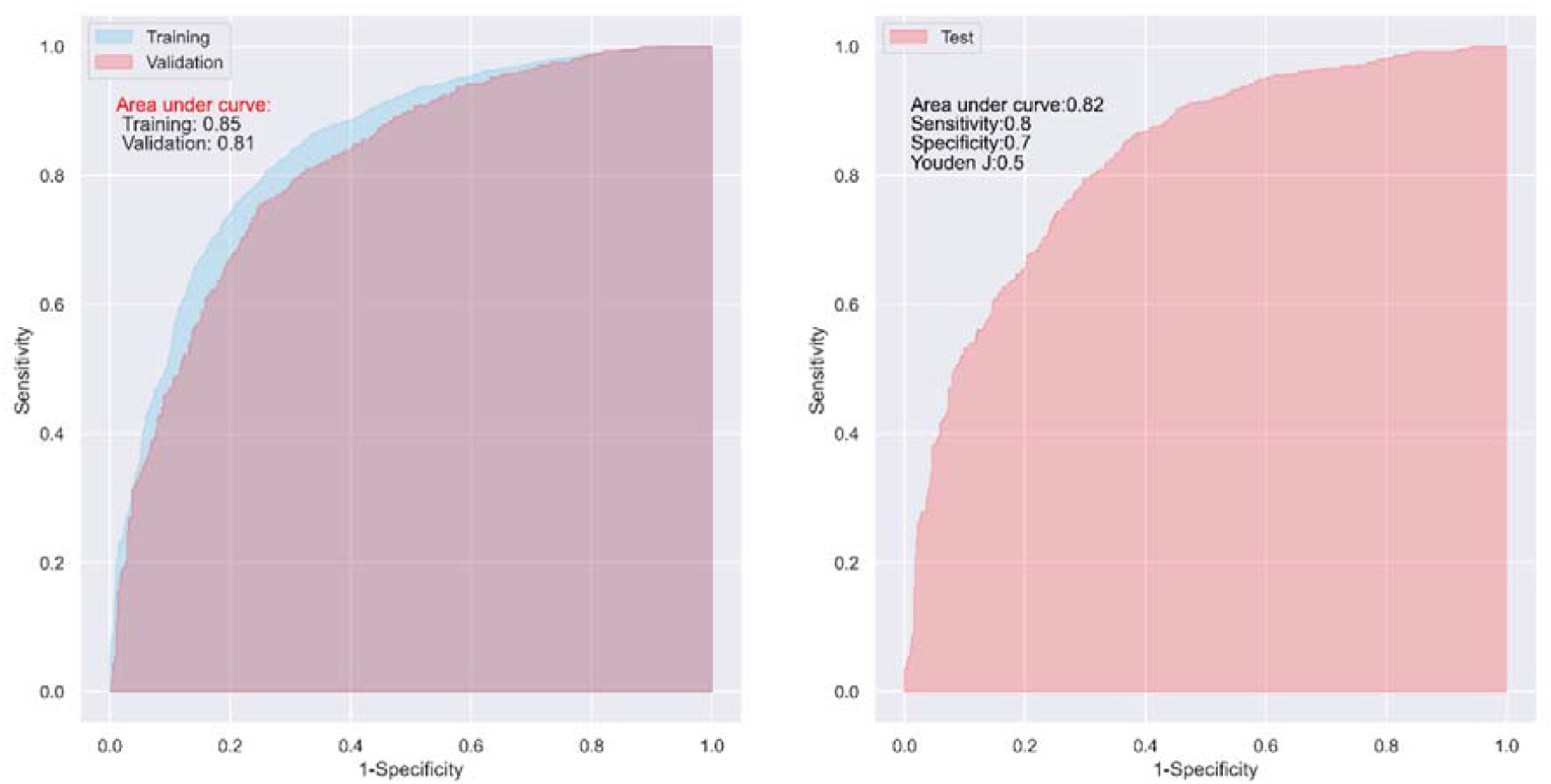
ROC curves of the CNN model in one of 100 trainings and validations (left), and tests (right) for complete remission

**Supplementary Figure S3.**
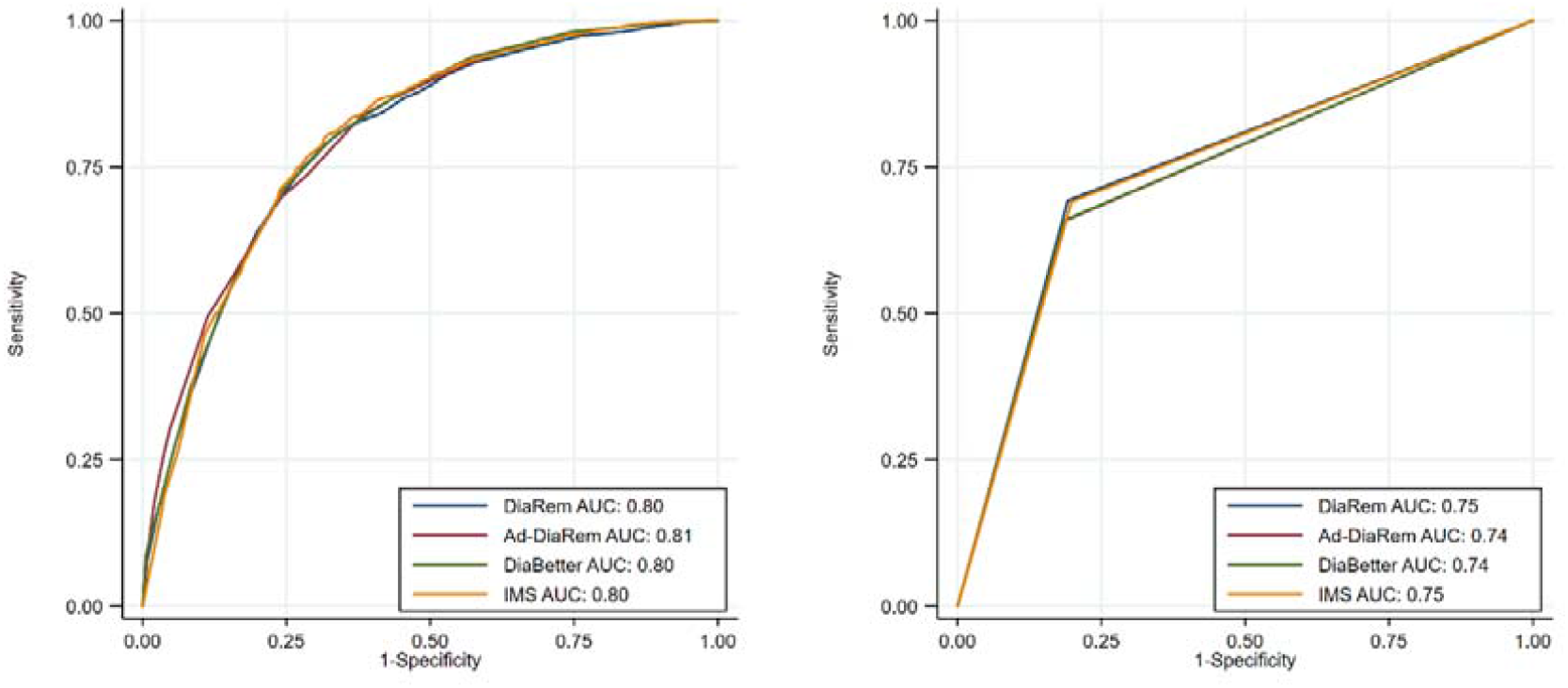
ROC curves of diabetes indices in one of 100 trainings (left) and tests (right) for complete remission.

**Supplementary Figure S4.**
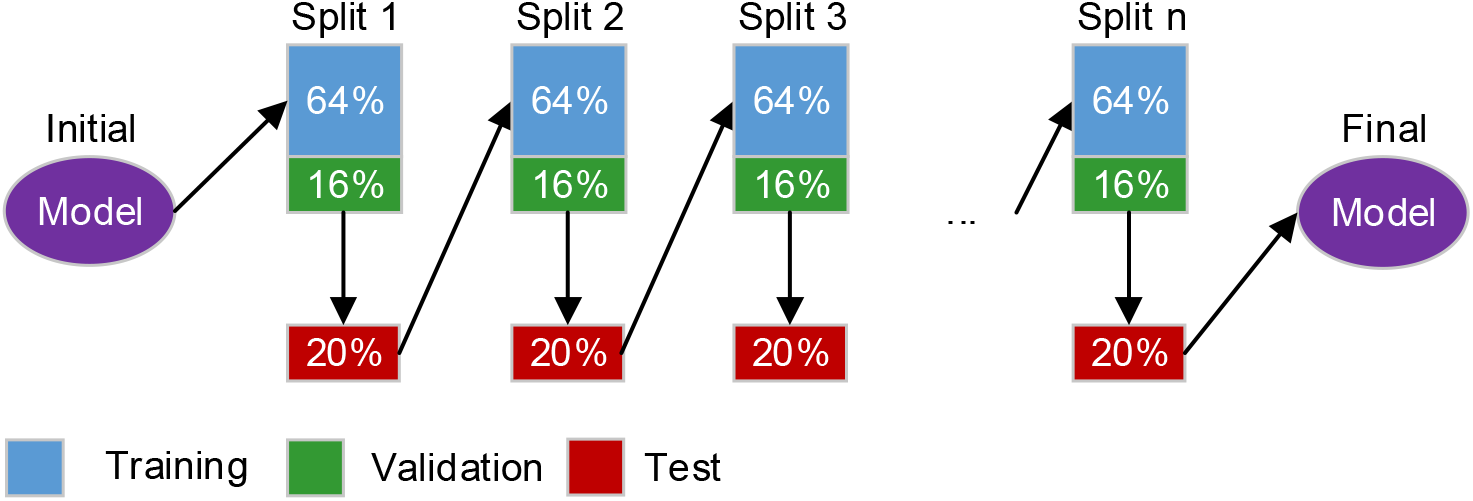
A sequentially model training process

**Supplementary Figure S5.**
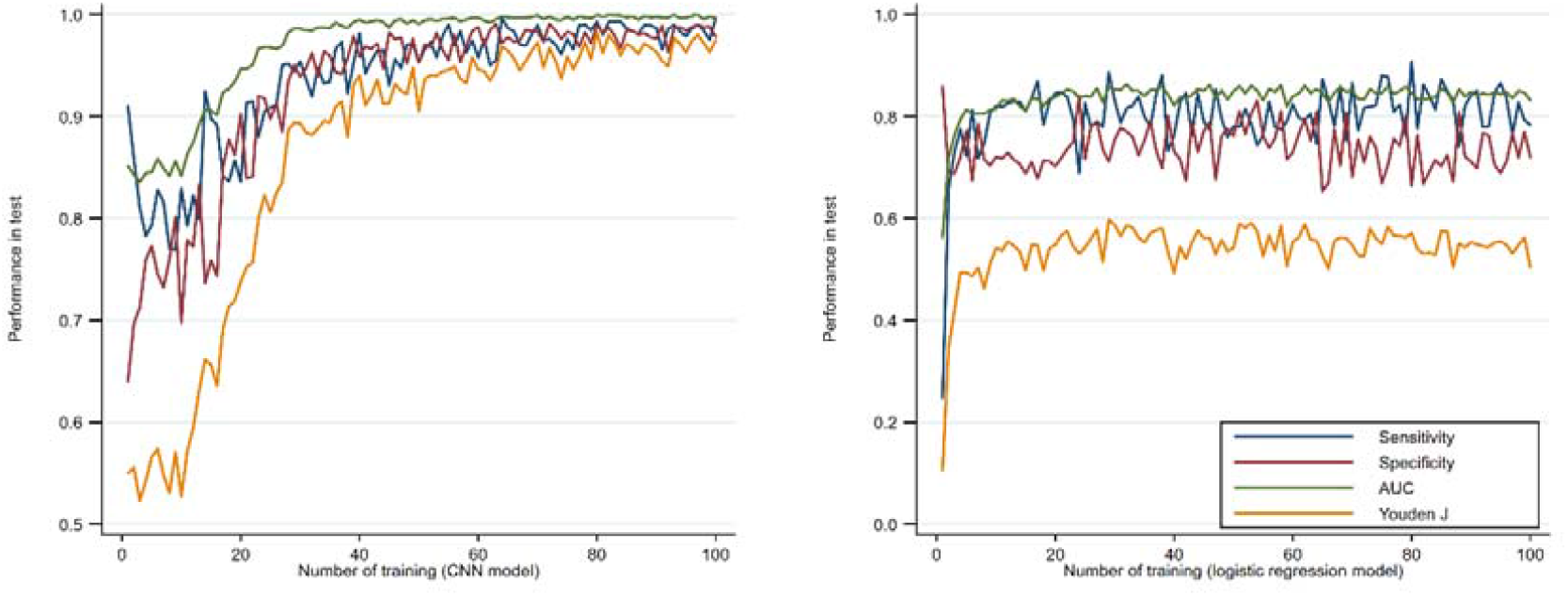
Predictive ability of the CNN model (left) and the logistic regression model (right) in the test dataset using the continuously training process

